# County-Level Structural Racism and Racial Disparities in Hypertensive Disorders of Pregnancy in Georgia

**DOI:** 10.64898/2026.09.05.26362353

**Authors:** Sheree L Boulet, Ran Zhang, Reem Abdelghany, Rachel Kienle, Kaitlyn K Stanhope, Jasmin Darville, Michael R Kramer, Sierra Carter

**Affiliations:** Department of Obstetrics and Gynecology, Chobanian & Avedisian School of Medicine, Boston University, Boston, MA; Department of Obstetrics and Gynecology, Boston Medical Center, Boston, MA; Department of Biostatistics & Epidemiology, School of Public Health, Boston University, Boston, MA; Department of Medicine, Emory University School of Medicine, Atlanta, GA; Center for Rural Health & Health Disparities, School of Medicine, Mercer University, Macon, GA; Department of Psychology, University of Georgia, Athens, GA

## Abstract

**Background:** Racialized disparities in hypertensive disorders of pregnancy persist, yet the role of interconnected structural determinants has not been fully explored. We evaluated the association between county-level indicators of structural racial discrimination (SRD) and Black-White disparities in HDP in Georgia.

**Methods:** We conducted exploratory factor analysis to identify latent factors of county-level SRD from 12 indicators representing historic racial violence, criminal justice, residential segregation, and political representation. We linked factor scores to 2020-2024 Georgia natality data and estimated associations between factor scores (tertiles) and Black-White differences in HDP rates.

**Results:** We identified two latent factors: Historic Racism and Racialized Polarization. HDP occurred in 15.5% of 206,438 Black births and 13.1% of 265,693 White births. Compared with the first tertile of Historic Racism factor scores, scores in the second (T2) and third (T3) tertiles were associated with increased Black-White HDP disparities (T2: 6.3, 95% confidence interval [CI]: –3.0-15.5; T3: 5.0, 95% CI: 5.4-15.4). Higher Racialized Polarization scores were associated with increased Black-White HDP disparities (T2: 9.1, 95% CI: –0.3-18.5; T3: 9.8, 95% CI: –1.5-21.2).

**Conclusions:** Structural racism contributes to racial disparities in HDP in Georgia, where historic oppression remains embedded in contemporary legal, political, and social systems.

## Introduction

Hypertensive disorders of pregnancy (HDP) constitute a major contributor to maternal morbidity and mortality in the United States and remain a leading cause of preventable obstetric complications.^1^ HDP, including chronic hypertension, gestational hypertension, preeclampsia, and eclampsia, complicate approximately 10% of pregnancies nationally^2,3^ and are associated with increased risks of stroke, organ failure, preterm birth, and long-term cardiovascular disease.^4–6^ Prior studies have documented a higher burden of HDP among Black women, alongside earlier onset and poorer postpartum cardiovascular trajectories.^7,8^

Structural racial discrimination (SRD) includes the structures, policies, and practices operating within social institutions that perpetuate unequal access to opportunities and resources by privileging some racial groups while oppressing others.^9^ SRD are fundamental drivers of racial inequities in maternal health, operating through interconnected social, political, and economic systems, including residential segregation, political exclusion, criminalization, and historical racial violence, each of which is spatially patterned and sustained over time.^10^ These systems shape chronic stress exposure, material deprivation, and healthcare access, all of which are biologically and socially relevant pathways for the development of HDP.^9,10^

Although a growing body of literature has examined the relationship between SRD and adverse birth outcomes,^10,11^ empirical evidence linking SRD to HDP remains limited. Most studies to date have focused on narrow domains of SRD, overlooked the interconnected nature of SRD measures, or relied on indices that assign equal weight to each indicator.^12–15^ In a prior study, we examined associations between 11 county-level indicators of SRD and disparities in severe maternal morbidity in Georgia, a state where the enduring legacy of one of the largest enslaved populations in the United States continues to shape contemporary maternal health outcomes.^16^ Building on this work, the current study used exploratory factor analysis to derive a reduced set of latent SRD factors from county-level indicators and estimate the association between SRD factors and racialized disparities in HDP.

## Methods

### Exploratory Factor Analysis

We operationalized county-level SRD using 12 indicators representing four domains outlined in our prior conceptual framework: historic racial violence, criminal justice, residential segregation, and political representation.^16^ Historic SRD was measured using the proportion of residents enslaved in 1860 and counts of documented lynchings between 1865 and 1950. Criminal justice inequities were represented by racial disparities in rates of felony incarcerations and counts of fatal police encounters. Residential SRD was captured using the Index of Concentration at the Extremes (ICE, using single-component measures of income and race and a multicomponent measure of race + income),^17^ the Dissimilarity Index,^18^ and Black-White disparities in proportions of residents living in rental housing. Political participation was assessed through racial differences in voter participation rates, the proportion of Black-presenting county elected officials, and disparities between the racial composition of county elected officials and constituents in the county.^19^ We transformed variables as needed and standardized all indicators using z-scores prior to factor extraction. Please see **Supplemental Table 1** for detailed descriptions, data sources, and calculation methods for each indicator. Please see the **Major Resources Table** in the Supplemental Materials for datasets.

We conducted initial exploratory factor analysis (EFA) with the minimum residual extraction method and oblique rotation (promax), using eigenvalues, scree plot inspection, and parallel analysis to guide factor retention. We iteratively evaluated the factor solution and removed indicators with high correlations (>.80), high communalities, or unstable cross-loadings to identify the most appropriate, parsimonious model. We used the Root Mean Square Error of Approximation (RMSEA), Root Mean Square of Residuals (RMSR), and Bayesian Information Criterion (BIC) to assess model fit. We evaluated the absolute values of loading scores and considered values >0.3 to be associated with the factor. Factor scores were computed using the Thurstone method. As the sample size was limited to the total number of counties in Georgia (n=159), we were unable to split the dataset for testing and validation. Therefore, we used Bayesian EFA to confirm the robustness of the traditional EFA results, assessing fit using the factor correlation posterior mean.^20^ We specified the Bayesian EFA model using the indicators retained from the traditional EFA model, using 5,000 Markov chain Monte Carlo (MCMC) iterations and restricting to no more than two latent factors, based on the results of the initial EFA. Given the suitability of Bayesian EFA for smaller samples, we used county-level factor scores generated from the Bayesian EFA in subsequent analyses.

## Spatial analysis

We analyzed live births to Georgia residents aged 15-49 years using restricted-use US natality data (2020-2024).^21^ To evaluate Black-White disparities, we restricted the study population to individuals identifying as non-Hispanic Black or non-Hispanic White and excluded records with missing data on chronic hypertension, gestational hypertension, or eclampsia. The unit of analysis was maternal county of residence.

We ascertained HDP from birth certificate data, defining HDP as report of pre-pregnancy (chronic) hypertension, gestational hypertension (including pregnancy-induced hypertension and preeclampsia), or eclampsia.^22^ We described distributions of individual-level characteristics of the study population, including maternal age at birth, pre-pregnancy body mass index (BMI, measured in kilograms/meter^2^), education, marital status, parity, insurance status at delivery, and prevalence of chronic and gestational diabetes. We defined rurality of maternal residence as a dichotomous measure, using Rural-Urban Continuum Codes of 4-9.^23^

We used Bayesian conditional autoregressive (CAR) Poisson models with Besag, York, and Mollié priors and queen adjacency to estimate county-specific, smoothed HDP rates for Black and White births, aggregated across the study period and adjusted for maternal age at birth. To minimize instability in county-level estimates, we excluded counties with <50 births to Black or White individuals during the study period. We calculated risk differences by subtracting age-adjusted HDP rates among Black births from those among White births. We categorized county-level factor scores into tertiles and used Bayesian CAR Poisson models to estimate adjusted risk ratios (aRR) and 95% confidence intervals (CI) for the association between the identified factors and HDP adjusting for age, rurality, and Black:White voting ratios (initially considered in the EFA but retained as a confounder as it loaded as a distinct factor). To evaluate the association between the SRD factors and the magnitude of Black-White disparities in HDP, we used Bayesian CAR linear models to estimate the association between tertiles of factor scores and the difference between the Black-White rate differences in HDP, adjusting for age, rurality and Black:White voting ratios. No data were missing for the county-level analyses.

We also conducted sensitivity analyses to explore use of different thresholds for characterizing the factor scores (quintiles) and evaluating factor scores as continuous predictors. For the quintiles, we applied the same method as the tertile models and categorized the scores into five groups, with the first quintile as the referent. For the continuous factor scores, we fit Bayesian linear and nonlinear spline models using a second-order random walk and assumed Gaussian distributions. We compared fit using the Deviance Information Criterion (DIC) and Watanabe-Akaike Information Criterion (WAIC).

We conducted all analyses using SAS version 9.4 and R version 4.5.0. We considered p-values <.05 statistically significant for the non-Bayesian comparisons. Our Institutional Review Board determined that this study was not human subjects research.

## Results

### Exploratory Factor Analysis

Of the 12 indicators initially considered in the EFA, seven were retained and subsequently classified into two factors reflecting related but distinct dimensions of SRD, *Historic Racism* and *Racialized Polarization* (**Table 1**). We removed ICE race-income, proportion of Black-presenting county elected officials, and counts of fatal police encounters due to evidence of strong multicollinearity. We also excluded county-level incarceration rates because they did not demonstrate meaningful factor loadings on either factor. Although parallel analysis suggested a potential three-factor solution, with the Black:White voting ratio loading as a distinct factor, we opted for a more parsimonious two-factor solution and retained the Black:White voting ratio as a separate predictor in subsequent models. See **Supplemental Table 2** for full description of traditional EFA factor loadings and measures of communality, uniqueness, and complexity. In the final model, the Historic Racism factor showed the strongest loading for the percentage of residents enslaved in 1860 (loading=0.84), with moderate contributions from ICE race (0.53), lynching counts (0.45), and the Black-White rental housing ratio (0.32). The Racialized Polarization factor was dominated by ICE income (1.04), with additional loadings from differences in racial representation of county elected officials (0.36) and the Dissimilarity Index (–0.34). Model fit indices indicated moderate fit given the small sample size of 159 counties (RMSEA = 0.103, 90% CI: 0.052–0.157; RMSR = 0.06; off-diagonal fit = 0.95).

**Table 1.** Final factor loadings for the structural racial discrimination indicators.

| Indicator | Traditional EFA<sup>1</sup> |  | Bayesian EFA<sup>2</sup> |  |
| --- | --- | --- | --- | --- |
|  | Historic Racism | Racialized Polarization | Historic Racism | Racialized Polarization |
| Index of concentration at the extremes - income | -0.26 | 1.04 |  | 0.64 |
| Index of concentration at the extremes - race | 0.53 | 0.15 | 0.69 |  |
| Percentage of residents enslaved in 1860 | 0.84 | -0.14 | 0.66 |  |
| Count of documented lynchings 1865-1950 | 0.45 | -0.04 | 0.42 |  |
| Difference in racial representation between residents and county officials | 0.05 | 0.36 |  | 0.48 |
| Black: White rental ratio | 0.32 | 0.02 | 0.35 |  |
| Dissimilarity index | -0.19 | -0.34 |  | -0.57 |

Abbreviations: EFA, exploratory factor analysis <sup>1</sup>RMSEA 0.103 (90% CI: 0.052-0.157), BIC -19.0 <sup>2</sup>Factor correlation posterior mean 0.524

The Bayesian EFA largely confirmed the results of traditional EFA (posterior mean factor correlation of 0.52), with some differences in loadings. Within the Historic Racism factor, ICE race had the highest loading score (0.69), followed by the percentage of residents enslaved in 1860 (0.66), lynching counts (0.42) and Black-White rental housing ratio (0.35). For the Racialized Polarization factor, ICE income had the highest loading (0.64), followed by the Dissimilarity Index (–0.57) and differences in racial representation of county elected officials (0.48).

### Study Population

A total of 623,647 births to Georgia residents aged 15-49 years occurred during 2020-2024. We excluded 151,516 births to individuals who did not identify as non-Hispanic Black or non-Hispanic White and 558 records with missing data on HDP. The final analytic sample was composed of 471,573 births, including 206,191 (43.7%) births to non-Hispanic Black women and 265,382 (56.3%) births to non-Hispanic White women. All records included had valid data on maternal county of residence.

Most women delivered between ages 20-34 years (77.8%). (**Table 2**) Black women were less likely than White women to have a college degree (22.9% vs. 41.9%), be married (29.1% vs. 72.9%), or reside in a rural area (12.9% vs 22.1%) and more likely to have Medicaid insurance at delivery (64.2% vs. 32.9%). Black women had higher prevalence of obesity (41% vs. 30.2%) and chronic diabetes (1.5% vs. 0.9%) than White women but lower prevalence of gestational diabetes (5.6% vs. 6.4%). HDP occurred more frequently among Black births than White births (15.5% vs. 13.1%).

**Table 2.** Distribution of demographic and clinical characteristics of Georgia births, 2020-2024.

| **Characteristic** | **Total**<br>**N (%)** | **Non-Hispanic**<br>**Black**<br>**N (%)** | **Non-Hispanic**<br>**White**<br>**N (%)** | **p-value** |
| --- | --- | --- | --- | --- |
| *N* | 471,573 | 206,191 (43.7) | 265,382 (56.3) |  |
| **Age (Years)** |  |  |  | <0.001 |
| 15-19 | 22,068 (4.7) | 12,578 (6.1) | 9,490 (3.6) |  |
| 20-34 | 367,011 (77.8) | 157,115 (76.2) | 209,896 (79.1) |  |
| 35-49 | 82,494 (17.5) | 36,498 (17.7) | 45,996 (17.3) |  |
| **Education** |  |  |  | <0.001 |
| Less than high school | 38,354 (8.2) | 18,476 (9.0) | 19,878 (7.5) |  |
| High school | 151,096 (32.1) | 83,254 (40.5) | 67,842 (25.6) |  |
| Some college | 122,948 (26.1) | 56,848 (27.6) | 66,100 (25.0) |  |
| College graduate | 158,034 (33.6) | 47,053 (22.9) | 110,981 (41.9) |  |
| Missing | 1,141 | 560 | 581 |  |
| **Married** | 253,382 (53.7) | 59,927 (29.1) | 193,455 (72.9) | <0.001 |
| **Parity** |  |  |  | <0.001 |
| 0 | 187,023 (39.7) | 76,663 (37.2) | 110,360 (41.7) |  |
| 1 | 145,526 (30.9) | 57,237 (27.8) | 88,289 (33.3) |  |
| 2+ | 138,197 (29.4) | 71,917 (34.9) | 66,280 (25.0) |  |
| Missing | 827 | 374 | 453 |  |
| **Insurance type** |  |  |  | <0.001 |
| Private | 209,324 (44.4) | 58,647 (28.5) | 150,677 (56.8) |  |
| Medicaid | 219,553 (46.6) | 132,260 (64.2) | 87,293 (32.9) |  |
| Self-pay | 13,375 (2.8) | 5,528 (2.7) | 7,847 (3.0) |  |
| Other | 28,805 (6.1) | 9,529 (4.6) | 19,276 (7.3) |  |
| Missing | 516 | 227 | 289 |  |
| **Rural residence** | 85,284 (18.1) | 26,669 (12.9) | 58,615 (22.1) | <0.001 |
| **Body mass index (kg/m2)** |  |  |  | <0.001 |
| Underweight | 13,083 (2.8) | 5,383 (2.6) | 7,700 (2.9) |  |
| Normal | 168,468 (35.9) | 60,979 (29.7) | 107,489 (40.7) |  |
| Overweight | 124,104 (26.4) | 54,786 (26.7) | 69,318 (26.2) |  |
| Obese | 163,876 (34.9) | 84,138 (41.0) | 79,738 (30.2) |  |
| Missing | 2042 | 905 | 1137 |  |
| **Pregnancy complications** |  |  |  |  |
| Chronic diabetes | 5,319 (1.1) | 2,994 (1.5) | 2,325 (0.9) | <0.001 |
| Gestational diabetes | 28,673 (6.1) | 11,593 (5.6) | 17,080 (6.4) | <0.001 |
| Hypertensive disorder of pregnancy | 66,974 (14.2) | 32,038 (15.5)<sup>1</sup> | 34,936 (13.1) | <0.001 |
| Chronic hypertension | 17,255 (3.7) | 10,099 (4.9) | 7,156 (2.7) | <0.001 |
| Gestational hypertension | 48,933 (10.4) | 21,565 (10.5) | 27,368 (10.3) | 0.10 |
| Eclampsia | 787 (0.2) | 375 (0.2) | 412 (0.2) | 0.03 |

<sup>1</sup>Includes one birth with gestational hypertension and eclampsia reported

### Spatial Distribution of HDP and SRD Factors

Across the 136 Georgia counties included in spatial analyses, the average age-adjusted HDP rate was 164.3 per 1,000 Black and White births, with higher rates concentrated in central and southern counties (**Figure 1**). The average county-level age-adjusted HDP rate was 181.4 per 1000 births among Black women and 152.5 per 1,000 births among White women. Spatial patterns of HDP rates were similar for both groups. The average Black-White risk difference was 28.9 per 1,000 births, and counties with the largest disparities were dispersed across the state.

**Figure 1.**
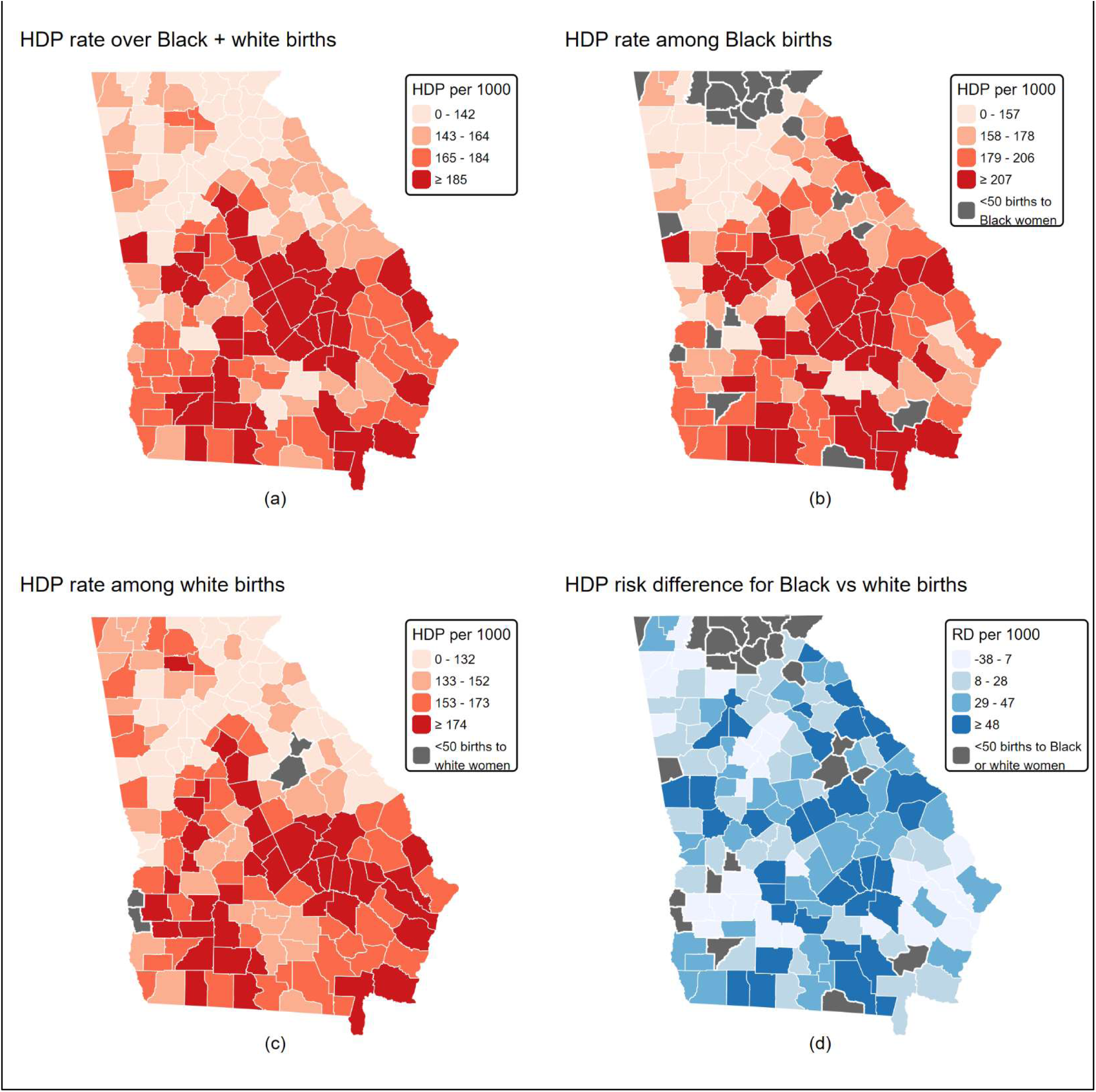
County-level distribution of hypertensive disorders of pregnancy (HDP) rates and racial disparities across Georgia.

As depicted in **Supplemental Figure 1**, there were fewer Black births in the northernmost part of Georgia, with the highest proportions occurring in metropolitan areas, including Atlanta, Augusta, Columbus, and Savannah. The geographic distributions of the Historic Racism and Racialized Polarization factor scores differed across counties. Counties in the Atlanta metropolitan area generally had higher Historic Racism factor scores but lower Racialized Polarization factor scores.

Among the 136 Georgia counties with ≥50 births to Black or White women during the study period, age-adjusted HDP rates tended to increase as Historic Racism factor scores increased for both Black and White births (**Table 3**). Among Black births, the adjusted rate ratios (aRRs) for the association between tertiles of Historic Racism and HDP were 1.15 (95% CI: 1.03-1.27) for births in counties with factor scores in second tertile (T2) and 1.07 (95% CI: 0.96-1.20) for births in counties with scores in third tertile (T3), relative to the first tertile (T1). Among White births, corresponding aRRs were 1.06 (95% CI: 0.95-1.19) and 1.00 (95% CI: 0.87-1.14) for T2 and T3, respectively. The Black-White rate difference increased by 6.30 per 1,000 births (95% CI: –3.00-15.50) in T2 and 5.00 per 1,000 (95% CI: –5.40-15.40) in T3, compared with T1.

**Table 3.**
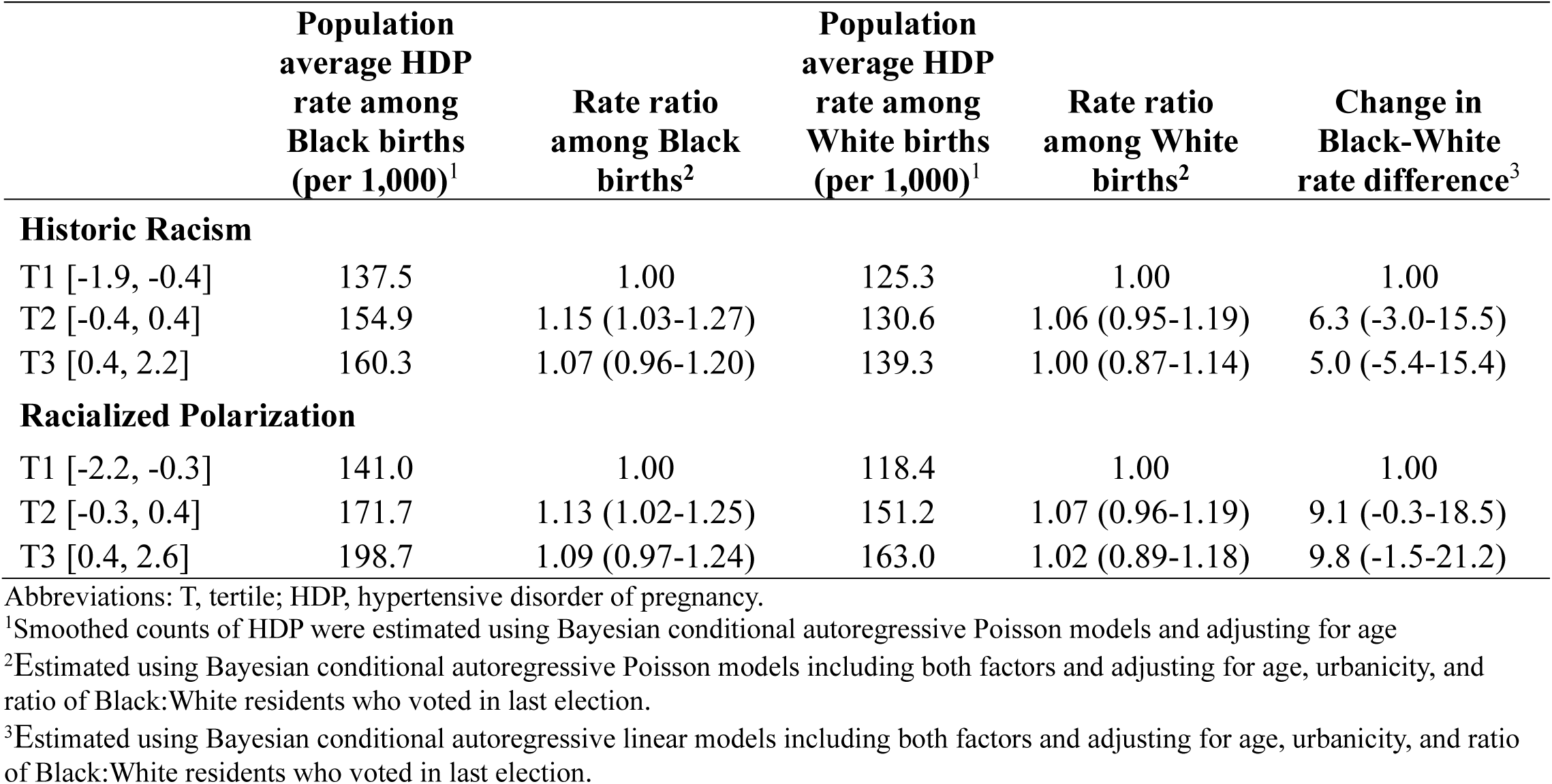
Association between structural racial discrimination factors and hypertensive disorders of pregnancy, among 136 Georgia counties with 50 or more births to Black or White women, 2020-2024.

Similar patterns were observed for the Racialized Polarization factor. Among Black births, residing in counties with Racialized Polarization factor scores in the upper tertiles was associated with increased HDP rates compared with first tertile (T2: aRR 1.13, 95% CI: 1.02-1.25; T3: aRR 1.09, 95% CI: 0.97-1.24). Among White births, the corresponding associations were weaker (T2: aRR 1.07, 95% CI: 0.96-1.19; T3: aRR 1.02, 95% CI: 0.89-1.18). The Black-White rate difference was consistent across T2 and T3 (9.1 per 1,000, 95% CI: –0.3 to 18.5 and 9.8 per 1,000 births, 95% CI: –1.5 to 21.2, respectively), relative to T1.

#### Sensitivity Analyses

Sensitivity analyses using quintiles of the factor scores yielded findings consistent with the primary tertile classification (**Supplemental Table 3**). The associations between SRD factor scores and HDP appeared to strengthen at the third and fourth quintiles for both Black and White births, suggesting a possible threshold effect.

## Discussion

Using twelve county-level indicators of SRD in Georgia, we identified two latent constructs, Historic Racism and Racialized Polarization, that captured dimensions of racial and economic segregation, the legacy of enslavement and lynching, and disparities in political representation. While both factors were independently associated with racialized differences in HDP, the excess risk among Black births remained relatively stable across the upper tertiles of each factor, suggesting that even moderate levels of exposure to SRD may contribute to HDP disparities. Notably, the adjusted magnitude of HDP rate differences was greater for the Racialized Polarization construct than Historic Racism (approximately 9 per 1000 vs 5 per 1000, respectively), indicating that contemporary structural conditions may be more closely linked to current inequities in maternal cardiovascular risk when historical structural conditions are accounted for.

Our findings are consistent with previous studies documenting associations between HDP risk and measures of racial and economic segregation, the Social Vulnerability Index, the Area Deprivation Index, and ICE income.^12,13,15,24^ Our work extends this literature by moving beyond single-domain or equally weighted composite measures, aligning with recent calls for theoretically grounded approaches to conceptualize and measure SRD and its impact on health.^25,26^ By applying EFA to a broad set of indicators, we sought to capture the interconnected and multidimensional nature of SRD through a data-driven approach that allows latent constructs to emerge empirically instead of imposing a priori assumptions regarding their structure.^27^

The distinction between Historic Racism and Racialized Polarization provides important insight into how structurally embedded and temporally distinct processes of racial stratification may shape maternal cardiovascular health within Georgia’s county-level contexts. SRD represents a set of institutional and historical processes that systematically produce racial inequities in exposure to risk and access to resources across domains such as residential stratification, economic inequality, political exclusion, and criminalization.^9^ In Georgia, these histories are unevenly distributed, with many counties in the Black Belt and parts of southern Georgia exhibiting persistent markers of historical racial violence and structural exclusion.^28,29^ Prior evidence has shown that counties with higher historical lynching frequency continue to experience worse population health, including lower life expectancy, underscoring the intergenerational persistence of these structural conditions.^30^

In contrast, the Racialized Polarization construct captures more contemporary patterns of economic stratification, residential separation, and political representation across Georgia counties. These dynamics are particularly evident in the contrast between rapidly expanding metropolitan regions, such as the Atlanta metropolitan area, and rural counties with persistent population loss and limited institutional resources.^31^ Such polarization reflects geographic separation and inequitable distribution of political power, economic investment, and institutional resources, creating conditions that may influence maternal cardiovascular health through chronic stress, access to high-quality care, and broader social conditions during pregnancy.^32^

Notably, the Dissimilarity Index loaded inversely on the Racialized Polarization factor, indicating that higher factor scores were associated with lower levels of racial segregation as measured by this index. Although initially counterintuitive, several explanations may account for this finding. First, counties may not represent the most appropriate geographic unit for measuring residential segregation using the Dissimilarity Index, which was developed primarily to assess patterns across metropolitan areas.^33^ Second, the Index measures the evenness of the distribution of two groups but does not account for isolation; therefore, counties with relatively balanced racial compositions may still experience substantial separation across local contexts.^33,34^ As such, when other measures (e.g., ICE) are accounted for, the Index may become protective as the lack of evenness may offer some degree of collective protection from daily discrimination. Moreover, residential integration does not always imply equitable access to resources, and systemic inequalities can persist even in integrated areas. This is particularly relevant in the context of gentrification and its potential to amplify health disparities. The inverse association may also reflect limitations of the Dissimilarity Index in increasingly diverse areas. Because the measure assesses the distribution of only two groups, it may not adequately capture how the presence and spatial distribution of other racial and ethnic populations shape access to opportunities and resources. Lastly, the Racialized Polarization factor included a measure of racial incongruence between a county’s elected officials and constituents.^19^ It is possible that some highly segregated counties contained geographically concentrated minority communities with stronger political organization and representation, resulting in lower levels of political polarization despite greater residential segregation.

Our study is subject to several limitations. As an ecological analysis, our findings cannot be used to infer individual-level associations. HDP diagnoses were derived from birth certificate data and may be subject to misclassification or underreporting. Prior research has shown that birth certificate data may underestimate chronic hypertension in pregnancy.^35^ In addition, the exploratory factor structure was developed specifically within Georgia and may not generalize to other states with different historical and demographic contexts.

### Perspectives

Structural racism contributes to racial disparities in HDP in Georgia, where historic practices remain embedded in contemporary legal, political, and social systems that continue to shape maternal health. Our findings underscore the importance of addressing structural determinants of maternal health and suggest that reducing maternal cardiovascular disparities will require interventions that extend beyond individual-level clinical care to include sustained investments in under-resourced areas, improved maternal healthcare infrastructure and access, and policies that promote more equitable political representation and resource distribution.

Moreover, this study supports the use of multidimensional SRD measures to more fully capture the complexity of racism and highlights the need for comparative and multi-state studies to advance understanding of how structural racism shapes maternal health outcomes across contexts and over time.

### Novelty and Relevance

1. What is new?
  - We used exploratory factor analysis to identify underlying relationships (known as factors) among twelve variables that measure structural racism at the county level.
  - We calculated factor scores for each county that measured the degree of structural racial discrimination that was present.
  - We examined the association between the factor scores and differences in rates of hypertensive disorders for Black women compared with White women.
2. What is relevant?
  - We identified two county-level factors, Historic Racism and Racialized Polarization, that included a total of seven structural racism indicators.
  - We found that differences between rates of hypertensive disorders for Black versus White women were higher in counties with more structural racism compared with counties with less structural racism.
3. Clinical/pathophysiological implications?
  - Structural racism may contribute to racial disparities in rates of hypertensive disorders of pregnancy.
  - Efforts to improve equitable access to high-quality healthcare and health-promoting resources are needed to reduce racial disparities in hypertension before, during, and after pregnancy.

## Ethics approval

This study was approved by the Boston University Medical Campus Institutional Review Board.

## Data Availability

The structural racism data are available on our website. We cannot share the NCHS restricted use vital statistics data, but they are available upon request from NCHS.

https://www.bumc.bu.edu/obgyn/files/2026/08/GA-SRD-data-dictionary.xlsx

https://www.bumc.bu.edu/obgyn/files/2026/08/finalsrddata.csv

## Acknowledgements

Research reported in this publication was supported by the Eunice Kennedy Shriver National Institute of Child Health & Human Development of the National Institutes of Health (5R01HD109005).

## References

1. Gestational Hypertension and Preeclampsia: ACOG Practice Bulletin, Number 222. Obstet Gynecol. 2020;135(6):e237–e260. doi:10.1097/AOG.0000000000003891

2. Lam EL, Khan SS, Shah NS. Hypertensive Disorders of Pregnancy in the United States, 2016-2024. J Am Coll Cardiol. Published online May 13, 2026:S0735-1097(26)06133-4. doi:10.1016/j.jacc.2026.03.154

3. Bruno AM, Allshouse AA, Metz TD, Theilen LH. Trends in Hypertensive Disorders of Pregnancy in the United States From 1989 to 2020. Obstetrics & Gynecology. 2022;140(1):83–86. doi:10.1097/AOG.0000000000004824

4. Brouwers L, van der Meiden-van Roest AJ, Savelkoul C, et al. Recurrence of pre-eclampsia and the risk of future hypertension and cardiovascular disease: a systematic review and meta-analysis. BJOG. 2018;125(13):1642–1654. doi:10.1111/1471-0528.15394

5. Brohan MP, Daly FP, Kelly L, et al. Hypertensive disorders of pregnancy and long-term risk of maternal stroke-a systematic review and meta-analysis. Am J Obstet Gynecol. 2023;229(3):248–268. doi:10.1016/j.ajog.2023.03.034

6. Battarbee AN, Sinkey RG, Harper LM, Oparil S, Tita ATN. Chronic hypertension in pregnancy. American Journal of Obstetrics and Gynecology. 2020;222(6):532–541. doi:10.1016/j.ajog.2019.11.1243

7. Hailu EM, Carmichael SL, Snowden JM, Lyndon A, Main E, Mujahid MS. Trends and Racial and Ethnic Disparities in Maternal Cardiovascular Health in California. JAHA. 2025;14(19):e039295. doi:10.1161/JAHA.124.039295

8. Bond RM, Bello NA, Ansong A, Ferdinand KC. Public health and system approach in eliminating disparities in hypertensive disorders and cardiovascular outcomes in non-Hispanic Black women across the pregnancy life course. American Heart Journal Plus: Cardiology Research and Practice. 2024;46:100445. doi:10.1016/j.ahjo.2024.100445

9. Williams DR, Lawrence JA, Davis BA. Racism and Health: Evidence and Needed Research. Annu Rev Public Health. 2019;40(1):105–125. doi:10.1146/annurev-publhealth-040218-043750

10. Hailu EM, Maddali SR, Snowden JM, Carmichael SL, Mujahid MS. Structural racism and adverse maternal health outcomes: A systematic review. Health Place. 2022;78:102923. doi:10.1016/j.healthplace.2022.102923

11. Montalmant KE, Ettinger AK. The Racial Disparities in Maternal Mortality and Impact of Structural Racism and Implicit Racial Bias on Pregnant Black Women: A Review of the Literature. J Racial and Ethnic Health Disparities. 2024;11(6):3658–3677. doi:10.1007/s40615-023-01816-x

12. Mayne SL, Yellayi D, Pool LR, Grobman WA, Kershaw KN. Racial Residential Segregation and Hypertensive Disorder of Pregnancy Among Women in Chicago: Analysis of Electronic Health Record Data. American Journal of Hypertension. 2018;31(11):1221–1227. doi:10.1093/ajh/hpy112

13. Huysman BC, Adler L, Cohen S, et al. Comparison of community-level deprivation indices and their association with adverse pregnancy outcomes. American Journal of Obstetrics & Gynecology MFM. 2026;8(6):101896. doi:10.1016/j.ajogmf.2026.101896

14. Stanhope KK, Adeyemi DI, Li T, Johnson T, Boulet SL. The relationship between the neighborhood built and social environment and hypertensive disorders of pregnancy: A scoping review. Ann Epidemiol. 2021;64:67–75. doi:10.1016/j.annepidem.2021.09.005

15. Hollenbach SJ, Thornburg LL, Glantz JC, Hill E. Associations Between Historically Redlined Districts and Racial Disparities in Current Obstetric Outcomes. JAMA Netw Open. 2021;4(9):e2126707. doi:10.1001/jamanetworkopen.2021.26707

16. Darville JA, Campbell K, Stanhope KK, Kendall A, Carter S, Kramer MR, Zhang R, Boulet SL. Using spatial Bayesian models to estimate associations between structural racial discrimination and disparities in severe maternal morbidity. Soc Sci Med. 2025;371:117932 10.1016/j.socscimed.2025.117932.

17. Krieger N, Waterman PD, Spasojevic J, Li W, Maduro G, Van Wye G. Public Health Monitoring of Privilege and Deprivation With the Index of Concentration at the Extremes. Am J Public Health. 2016;106(2):256–263. doi:10.2105/AJPH.2015.302955

18. Duncan OD, Duncan B. A Methodological Analysis of Segregation Indexes. American Sociological Review. 1955;20(2):210. doi:10.2307/2088328

19. Stanhope KK, Kapila P, Umerani A, et al. Political Representation and perinatal outcomes to Black, White, and Hispanic people in Georgia: A Cross-sectional Study. Ann Epidemiol. 2023;87:S1047-2797(23)00167-9. doi:10.1016/j.annepidem.2023.09.001

20. Conti G, Frühwirth-Schnatter S, Heckman JJ, Piatek R. Bayesian Exploratory Factor Analysis. J Econom. 2014;183(1):31–57. doi:10.1016/j.jeconom.2014.06.008

21. U.S. Centers for Disease Control and Prevention. Restricted-Use Vital Statistics Data. National Center for Health Statistics. August 24, 2023. Accessed May 20, 2026. https://www.cdc.gov/nchs/nvss/nvss-restricted-data.htm

22. National Center for Health Statistics. Guide to Completing the Facility Worksheets for the Certificate of Live Birth and Report of Fetal Death. 2019. Accessed March 27, 2026. https://www.cdc.gov/nchs/nvss/facility-worksheets-guide/14.htm

23. U.S. Department of Agriculture. Rural-Urban Continuum Codes | Economic Research Service. Accessed May 20, 2026. https://www.ers.usda.gov/data-products/rural-urban-continuum-codes

24. Francis B, Pearl M, Colen C, Shoben A, Sealy-Jefferson S. Racial and Economic Segregation Over the Life Course and Incident Hypertensive Disorders of Pregnancy Among Black Women in California. American Journal of Epidemiology. 2024;193(2):277–284. doi:10.1093/aje/kwad192

25. Brown TH, Homan P. Structural Racism and Health Stratification: Connecting Theory to Measurement. J Health Soc Behav. 2024;65(1):141–160. doi:10.1177/00221465231222924

26. Adkins-Jackson PB, Chantarat T, Bailey ZD, Ponce NA. Measuring Structural Racism: A Guide for Epidemiologists and Other Health Researchers. Am J Epidemiol. 2022;191(4):539–547. doi:10.1093/aje/kwab239

27. Watkins MW. Exploratory Factor Analysis: A Guide to Best Practice. Journal of Black Psychology. 2018;44(3):219–246. doi:10.1177/0095798418771807

28. Connor MA. Metropolitan Secession and the Space of Color-Blind Racism in Atlanta. Journal of Urban Affairs. 2015;37(4):436–461. doi:10.1111/juaf.12101

29. Gompers A, Lewis TT, Kramer MR. Structural racism and racial disparities in stroke mortality in the United States, 2021. Social Science & Medicine. 2025;366:117705. doi:10.1016/j.socscimed.2025.117705

30. Kihlström L, Kirby RS. We carry history within us: Anti-Black racism and the legacy of lynchings on life expectancy in the U.S. South. Health & Place. 2021;70:102618. doi:10.1016/j.healthplace.2021.102618

31. Georgia House of Representatives. House of Representatives Rural Development Council 2023 Overview and Recommendations. Georgia General Assembly. Accessed July 14, 2026. https://www.house.ga.gov/Documents/CommitteeDocuments/2023/Rural_Development_Council/House_Rural_Development_Council_2023_Final_Report.pdf

32. Bailey ZD, Krieger N, Agénor M, Graves J, Linos N, Bassett MT. Structural racism and health inequities in the USA: evidence and interventions. The Lancet. 2017;389(10077):1453–1463. doi:10.1016/S0140-6736(17)30569-X

33. White K, Haas JS, Williams DR. Elucidating the role of place in health care disparities: the example of racial/ethnic residential segregation. Health Serv Res. 2012;47(3 Pt 2):1278–1299. doi:10.1111/j.1475-6773.2012.01410.x

34. Kramer MR, Hogue CR. Is Segregation Bad for Your Health? Epidemiologic Reviews. 2009;31(1):178–194. doi:10.1093/epirev/mxp001

35. Delker E, Baer RJ, Chambers CD, Bandoli G. Identification of Chronic Hypertension in Pregnancy in Three Administrative Data Sources Among Medicaid-Funded Births in California. Pharmacoepidemiology and Drug Safety. 2024;33(12):e70059. doi:10.1002/pds.70059

